# Nucleocapsid and spike antibody responses post virologically confirmed SARS-CoV-2 infection: An observational analysis in the Virus Watch community cohort

**DOI:** 10.1101/2022.02.01.22270269

**Authors:** Annalan M D Navaratnam, Madhumita Shrotri, Vincent Nguyen, Isobel Braithwaite, Sarah Beale, Thomas E Byrne, Wing Lam Erica Fong, Ellen Fragaszy, Cyril Geismar, Susan Hoskins, Jana Kovar, Parth Patel, Alexei Yavlinsky, Anna Aryee, Alison Rodger, Andrew C Hayward, Robert W Aldridge, the Virus Watch Collaborative

**Affiliations:** Institute of Health Informatics, University College London, UK; Institute of Epidemiology and Health Care, University College London, London, UK; Department of Infectious Disease Epidemiology, London School of Hygiene and Tropical Medicine, Keppel Street, London, UK; Institute for Global Health, University College London, London, UK; Centre for Behaviour Change, University College London, London, UK; Department of Population, Policy and Practice, UCL Great Ormond Street Institute of Child Health, London, UK; Francis Crick Institute, London, UK; Health Protection and Influenza Research Group, Division of Epidemiology and Public Health, University of Nottingham School of Medicine, Nottingham, UK; University College London Hospital, London, UK; Department of Computer Science, University College London, London, UK; London Centre for Nanotechnology and Division of Medicine, London, University College London; SpaceTimeLab, Department of Civil, Environmental and Geomatic Engineering, University College London, London, UK

## Abstract

**Introduction:** Seroprevalence studies can provide a measure of cumulative incidence of SARS-CoV-2 infection, but a better understanding of antibody dynamics following infection is needed to assess longevity of detectability. Infection is characterised by detection of spike (anti-S) and nucleocapsid (anti-N) antibodies, whereas vaccination only stimulates anti-S. Consequently, in the context of a highly vaccinated population, presence of anti-N can be used as a marker of previous infection but waning over time may limit its use.

**Methods:** Adults aged ≥18 years old, from households enrolled in the Virus Watch prospective community cohort study in England and Wales, provided monthly capillary blood samples which were tested for anti-S and anti-N. Participants self-reported vaccination dates and past medical history. Prior polymerase chain reaction (PCR) swabs were obtained through Second Generation Surveillance System (SGSS) linkage data. Primary outcome variables were seropositivity (antibodies at or above the manufacturer’s cut-off for positivity) and total anti-N and anti-S levels after PCR confirmed infection. Outcomes were analysed by days since infection, self-reported demographic and clinical factors.

**Results:** A total of 13,802 eligible individuals, median age 63, provided 58,770 capillary blood samples. 537 of these had a prior positive PCR confirmed SARS-CoV-2 infection 0-269 days before the antibody sample date. 432 out of the 537 (80.44%) were anti-N positive and detection remained stable through-out follow-up. Median anti-N levels peaked between days 90 and 119 post PCR results and then began to decline. Logistic regression models, both univariable and multivariable, only showed higher odds of positive anti-N result between 0-269 days for 35-49 year olds, compared to 18-34 year olds. There is evidence of anti-N waning from 120 days onwards, with earlier waning for females and younger age categories.

**Discussion:** Approximately 4 in 5 participants with prior PCR-confirmed infection were anti-N positive, and this remained stable through follow-up for at least 269 days. However, median antibody levels began to decline from about 120 days post-infection. This suggests that anti-N have around 80% sensitivity for identifying previous COVID-19 infection and that this sensitivity is maintained through 269 days of follow up.

**Funding:** The research costs for the study have been supported by the MRC Grant Ref: MC_PC 19070 awarded to UCL on 30 March 2020 and MRC Grant Ref: MR/V028375/1 awarded on 17 August 2020. The study also received $15,000 of Facebook advertising credit to support a pilot social media recruitment campaign on 18th August 2020. The study also received funding from the UK Government Department of Health and Social Care’s Vaccine Evaluation Programme to provide monthly Thriva antibody tests to adult participants. This study was supported by the Wellcome Trust through a Wellcome Clinical Research Career Development Fellowship to RA [206602].

## Introduction

Antibodies produced following natural infection with severe acute respiratory syndrome coronavirus 2 (SARS-CoV-2), the virus which causes COVID-19, are known to provide some protection against reinfection for at least 6 months in the early stages of the pandemic.^1^ The proportion of infected individuals who are N-antibody (anti-N) and S-antibody (anti-S) positive and the stability of the antibody response over time are not well established.^2^ In the UK, surveillance has been largely through symptomatic testing with reverse transcriptase polymerase chain reaction (PCR) assays or asymptomatic testing through Lateral Flow Device (LFD) tests. In large scale population surveys for the Office for National Statistics (ONS) COVID Infection Study and the REal-time Assessment of Community Transmission (REACT) study and UK Health Security Agency (UKHSA) (formerly Public Health England) blood donation surveys, monitoring via seroprevalence is being carried out.^3–5^ Meta-analyses of the proportion of infections that are asymptomatic show approximately one third of cases do not develop symptoms at any point during acute infection.^6^

Seroprevalence studies can provide a measure of cumulative incidence that accounts for asymptomatic infections, but more information on antibody waning is needed to aid the interpretation of these studies. The widespread use of COVID-19 vaccines that only stimulate anti-S means that distinguishing antibodies derived from natural infection from those derived through vaccination requires measurement of both anti-N and anti-S.^7^ A challenge with seroprevalence studies is that it is still unclear how long anti-S and anti-N remain in circulation post-infection. The duration of which antibodies are detectable can also inform modelling approaches guiding the pandemic response, especially in countries where vaccine roll-out is in early stages. Much of the current evidence on antibody response or duration of detection is focused on specific occupational or institutional sub-groups (e.g healthcare, university, nursing homes).^8,9^ Assessing longer-term antibody responses across the population is critical to evaluate immune protection at the population level. Furthermore, anti-S are produced in response to both vaccination or previous infection and are therefore not an accurate measure of previous infection in countries that have rolled out widespread COVID vaccination programmes. Anti-N, which is only produced in response to SARS-CoV-2 infection, may be a reliable option for serosurveillance, but little is known about the timeline of seroconversion nor duration of detectable levels of antibodies.

To improve our understanding of the longevity of anti-N and anti-S responses over time, we evaluated both antibody detection and titres to establish factors that contribute to seropositivity and waning post infection, defined by previous positive PCR. Specifically, we aimed to investigate:

1. The proportion of individuals who are anti-N positive, within 269 days of PCR confirmed infection, and associated demographic and clinical characteristics.
2. Anti-N and anti-S detection and titres from 0-540 days since infection.
3. Comparison of anti-N and anti-S detection and titres based on infection, vaccination or both.

## Methods

### Study design and setting

The Virus Watch study is a household community cohort of acute respiratory infections in England and Wales that started recruitment in June 2020.^10^ As of 31 August 2021, 50,773 participants were recruited using a range of methods including post, social media and SMS messages and letters from their General Practice. Participants provided information on age, sex, ethnicity, household information (e.g. number of household members, postcode) and medical history (e.g. underlying medical conditions, medication history). Participants were followed-up weekly by email with a link to a survey that captured information about vaccination status and SARS-CoV-2 infection. Between February and May 2021, invitations to participate in monthly antibody testing were sent to enrolled eligible households. Consenting participants provided capillary blood samples on a monthly basis.

### Samples

Capillary blood samples (400-600µL) were self-collected by participants using an at-home kit, manufactured by the company Thriva. ^11^ Completed kits were returned by participants using pre-paid envelopes and priority postage boxes to United Kingdom Accreditation Service (UKAS)-accredited laboratories, for serological testing using Roche’s Elecsys Anti-SARS-CoV-2 electrochemiluminescence assays targeting total immunoglobulin (Ig) to the Nucleocapsid (N) protein, or to the receptor binding domain in the S1 subunit of the Spike protein (S) (Roche Diagnostics, Basel, Switzerland). Results for anti-N were reported as numeric values in the form of a cut-off index (COI). The detection limit is determined by the ratio of the luminescence of the sample relative to the predefined negative threshold. This threshold was determined by calibrating against known negative samples.^12^ At the manufacturer-recommended seropositivity thresholds (≥1.0 cut-off index [COI] for N and ≥0.8 units per millilitre [U/ml] for S), the N assay has a sensitivity of 97□2%-99.5% and specificity of 99.8%, while the S assay has a sensitivity of 97.9%-98.8% and a specificity of 100% with high agreement between the assays for samples from previously infected individuals. ^13–15^ All samples processed between 24th February 2021 to 31st August 2021 were included in the analysis of anti-N and, depending on vaccination status, anti-S results. For anti-S levels, samples processed before the 1st July (with the exception of a 2 day pilot) had an upper limit of 250U/ml. After this date, if sample results exceeded the upper limit of the analytical measuring interval they underwent dilution with diluent universal of 1:10 up to 1:100, providing an upper limit of 25,000 U/ml. ^16^

### Covariates

Age, sex, ethnicity, underlying health conditions, body mass index (BMI) and whether participants were taking immunosuppressants were self-reported during study registration. Age was grouped into the following categories: 18-34, 35-49, 50-64, 65-79 and ≥80 years. Ethnicity data were grouped into Black, White, South Asian, Other Asian, Mixed categories and Other/missing. Sex was limited to male and female categories. Participants were asked to report specific health conditions, which were grouped as part of the analysis (further details in Appendix Table 1).

**Table 1:** Proportion of those with positive anti-N result, grouped by age, sex and vaccination status

|  | Anti-N Positive (n / N) | Percentage Positive (%) | 95% CI |
| --- | --- | --- | --- |
| **Age category** |  |  |  |
| 18-34 | 58 / 79 | 73.42% | (63.68, 83.16) |
| 35-49 | 121 / 143 | 84.62% | (78.7, 90.53) |
| 50-64 | 151 / 193 | 78.24% | (72.42, 84.06) |
| 65-79 | 100 / 120 | 83.33% | (76.66, 90) |
| 80+ | 2 / 2 | 100% | (100, 100) |
| **Sex** |  |  |  |
| Female | 253 / 324 | 78.09% | (73.58, 82.59) |
| Male | 177 / 211 | 83.89% | (78.93, 88.85) |
| Missing data | 2 / 2 | 100% | (100, 100) |

| **Vaccinated** |  |  |  |
| --- | --- | --- | --- |
| No | 101 / 116 | 87.07% | (80.96,<br>93.18) |
| Yes | 308 / 396 | 77.78% | (73.68,<br>81.87) |
| Missing data | 23 / 25 | 92% | (81.37,<br>102.63) |

The primary source of data was the Virus Watch dataset linked to the Second Generation Surveillance System (SGSS), which contains SARS-CoV-2 PCR test results. The linkage period for SGSS encompassed data from April 2020 until August 2021. The earliest positive test result was taken as the date for PCR-antibody time analysis. We assumed that participants who did not have a positive PCR result in the SGSS never had virologically confirmed SARS-CoV-2 infection, so were therefore excluded from analysis.

Vaccination status was collected through the weekly Virus Watch questionnaire. This question was added to the weekly surveys on 11th January 2021, where individuals reported dates of their vaccination. Vaccination type was recorded in data collection and used to establish national licensure date for each type of vaccine (e.g. Pfizer 2nd December 2020, AstraZeneca 30th December 2020 and Moderna 8th January 2021). Results were excluded from analysis if the reported vaccination date preceded the national licensure date or if a second dose was reported but not a first dose. This was specifically relevant for comparison of anti-S and anti-N responses post-infection among those who had not been vaccinated or provided a sample pre-vaccination. When analysing response to vaccination in those with no evidence of prior infection only samples with negative anti-N were included.

### Primary outcome(s)

The main outcome variable was detection of anti-N expressed as a binary variable (≥1.0 COI), and anti-N level, expressed as the semi-quantitative COI, and subsequently log-transformed to base 10.

### Secondary outcomes

Secondary outcome was comparison of seropositivity to anti-S and anti-N, and levels in relation to prior infection and vaccination.

### Participants

Within the Virus Watch cohort, eligible households were defined as having at least one adult aged 18 years and over, a valid England or Wales postcode, and a complete postal address registered.^10^ Individuals that were 18 years and over within eligible households could consent to participate through provision of valid, electronic consent. Individuals were included in this analysis if they had provided at least one finger prick blood sample. Evidence of prior infection was defined as seropositivity to the Nucleocapsid protein (COI ≥ 1.0) among vaccinated individuals, or seropositivity to either Spike (≥0.8 U/ml) or Nucleocapsid protein among unvaccinated individuals. Samples with void anti-N or void anti-S results were excluded from analyses.

### Analysis

Analyses were carried out using two approaches: at individual level and at sample level. At individual level, samples obtained between 0-269 days post positive SARS-CoV-2 PCR were aggregated, selecting the sample with the first positive anti-N result if there was more than one sample per participant. Individual level data was used to compare age, sex, ethnicity, body mass index (BMI) or underlying medical conditions and if taking immunosuppressants. Proportions of individuals positive for anti-N or anti-S were calculated with 95% confidence intervals. To calculate odds of seroconversion after infection, and explore which explanatory variables influence this, univariable and multivariable logistic regression models were created using individual-level data from those with a previous positive PCR result. The multivariable model was adjusted for age and sex at birth, when they were not the explanatory variable being investigated. The explanatory variables included age categories, sex at birth, conditions associated with severe COVID-19 infection, obesity and whether taking any immunosuppressants or not (see Appendix; Table 1 and Table 2).

For antibody levels, as they are not normally distributed, median values for each group were calculated with interquartile ranges. Due to the available assay platform and dilution capabilities, the dynamic range of the anti-S assay was limited to 0.4-250 U/ml for samples processed before between February and June 2021. Only samples that underwent further dilution, from July 2021 onwards, were included in anti-S levels analysis. As dilution capabilities did not affect the threshold for a positive result, all anti-S samples were included when calculating proportion of positivity. For analysis of antibody positivity and levels by days post PCR confirmed infection (e.g. 0-29, 30-59, 60-89, 90-119, 120-269 and 270+ days after a positive PCR), all samples were included. The proportion of anti-N positive samples, with 95% confidence intervals, and median antibody levels were calculated for each time period category. Anti-N positivity and levels over time were stratified by age groups (18-49 and 50+) and sex. Comparison of anti-N and anti-S positivity and levels were made for unvaccinated individuals or those who provided a sample before vaccination.

Comparison of anti-N and anti-S positive levels were made between those with prior infection only (no vaccination) and those who had both vaccination and prior infection. Further comparison was made on anti-S positivity over time for those with prior infection only (no vaccination) and prior vaccination only (no infection), using the date of PCR test or vaccination date as day 0. Antibody levels were not normally distributed, therefore non-parametric tests were used to compare groups: the Mann-Whitney U (Wilcoxon Rank Sum) test for two groups, and the Kruskal-Wallis test for more than two groups with Benjamini & Hochberg correction. For comparison of binary variables, the Chi-Square test was used. All analyses were carried out with R studio (R 4.0.5.) using packages: ‘tidyverse’, ‘ggplot2’, ‘flextable’ and ‘rstatix’.

## Results

15,534 participants were consented and eligible to take part in the study, of which 13,802 participants provided 58,770 valid anti-N samples (Figure 1). The median age of participants was 63 (IQR 46, 80), and of 13,765 with a recorded sex at baseline survey, 57.07% (n = 7,856) were female. 5,331 samples from 1,479 individuals were anti-N positive.

**Figure 1:**
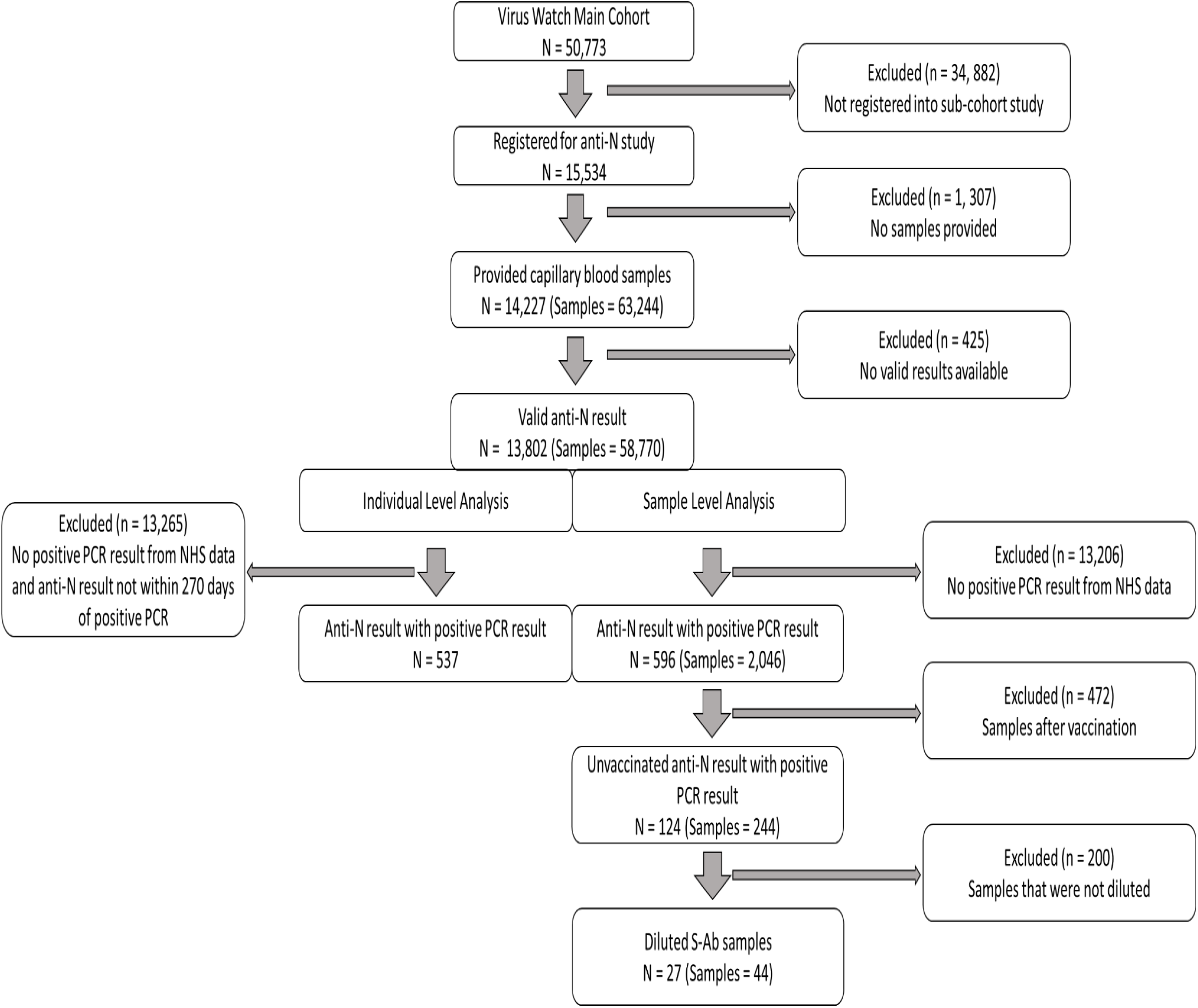
Study inclusion flow diagram

Of the 13,802 individuals with valid antibody results, 596 had a prior positive PCR result from the SGSS linkage data. 537 of those individuals had samples within 269 days of their PCR result, and were therefore included in the individual level analysis. 80.44% of individuals (432/537) were seropositive for anti-N after PCR confirmed infection. A univariable logistic regression model showed no evidence for a difference in being seropositive by age, sex or health condition. A multivariable logistic regression found higher odds of being seropositive if aged between 35-49 compared to age 18-34 (adjusted OR 1.98; p = 0.04), but no association was found for sex, obesity, conditions related to severe COVID19 infection or if taking any immunosuppressant (Appendix, Table 4).

We examined anti-N positivity and titres over time after infection, with PCR test date as day 0 (Figure 2). The minimum and maximum days post PCR were 1 day and 520 days (median 168.5 days, IQR 103.63 - 233.38), respectively. The proportion of samples with detectable levels of anti-N increased from 42.6% at days 0-29 post PCR (66/155, 95% CI 35-50) to 76% at 30-59 days (95/125, 95% CI 69-83), and then remained relatively stable at between 80-85% at 60-89, 90-119, 120-269 and 270+ days. Median anti-N titres reached a peak at around 60 COI between days 30 and 120, before beginning to decline. Patterns were similar in both sexes and different age groups although with some evidence of a faster decline in younger adults (18-49 years old) and an earlier peak in women than in men (Figure 2 and 3).

**Figure 2.**
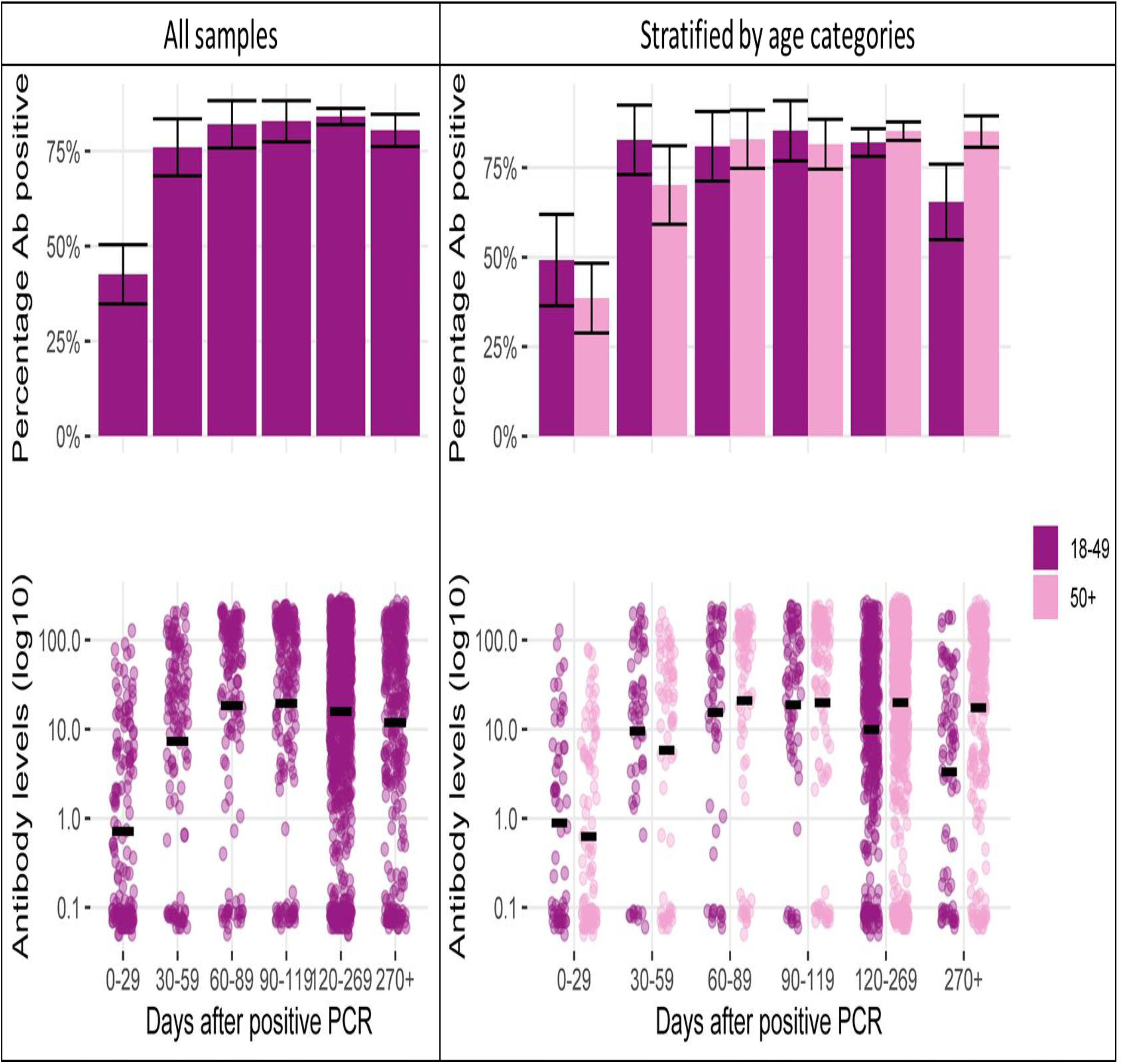
Anti-N percentage positivity and levels, stratified by days post positive PCR test and age. Notes: 95% confidence intervals represented by vertical cross bars and anti-N levels underwent Log10 transformation with bars representing the logarithmic mean.

**Figure 3:**
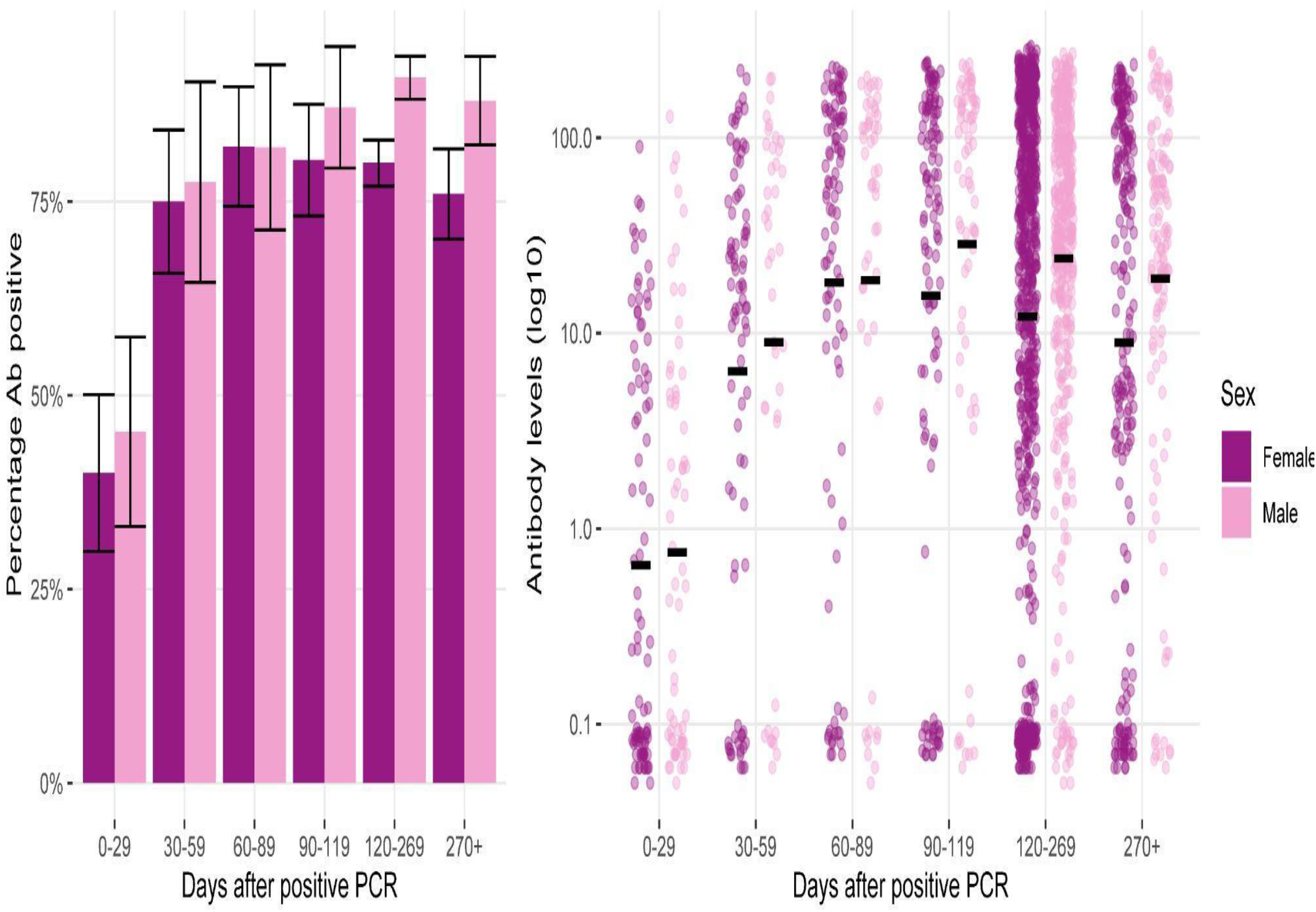
Anti-N percentage positivity and levels by sex and days after positive PCR Notes: 95% confidence intervals represented by vertical cross bars and anti-N levels underwent Log10 transformation with bars representing the logarithmic mean.

Of the 2,046 samples, 244 belonged to 124 individuals with only previous infection and no prior vaccination or only previous infection who were vaccinated after providing a capillary blood sample. The samples that underwent dilution (44 samples from 27 individuals), were included in anti-S level analysis. At 0-29 days, 60% (95% CI 30-90%) of samples had both detectable anti-S and anti-N but anti-S detectability had a sharper increase than anti-N and remained above 90% during the follow up period (Figure 4).

**Figure 4:**
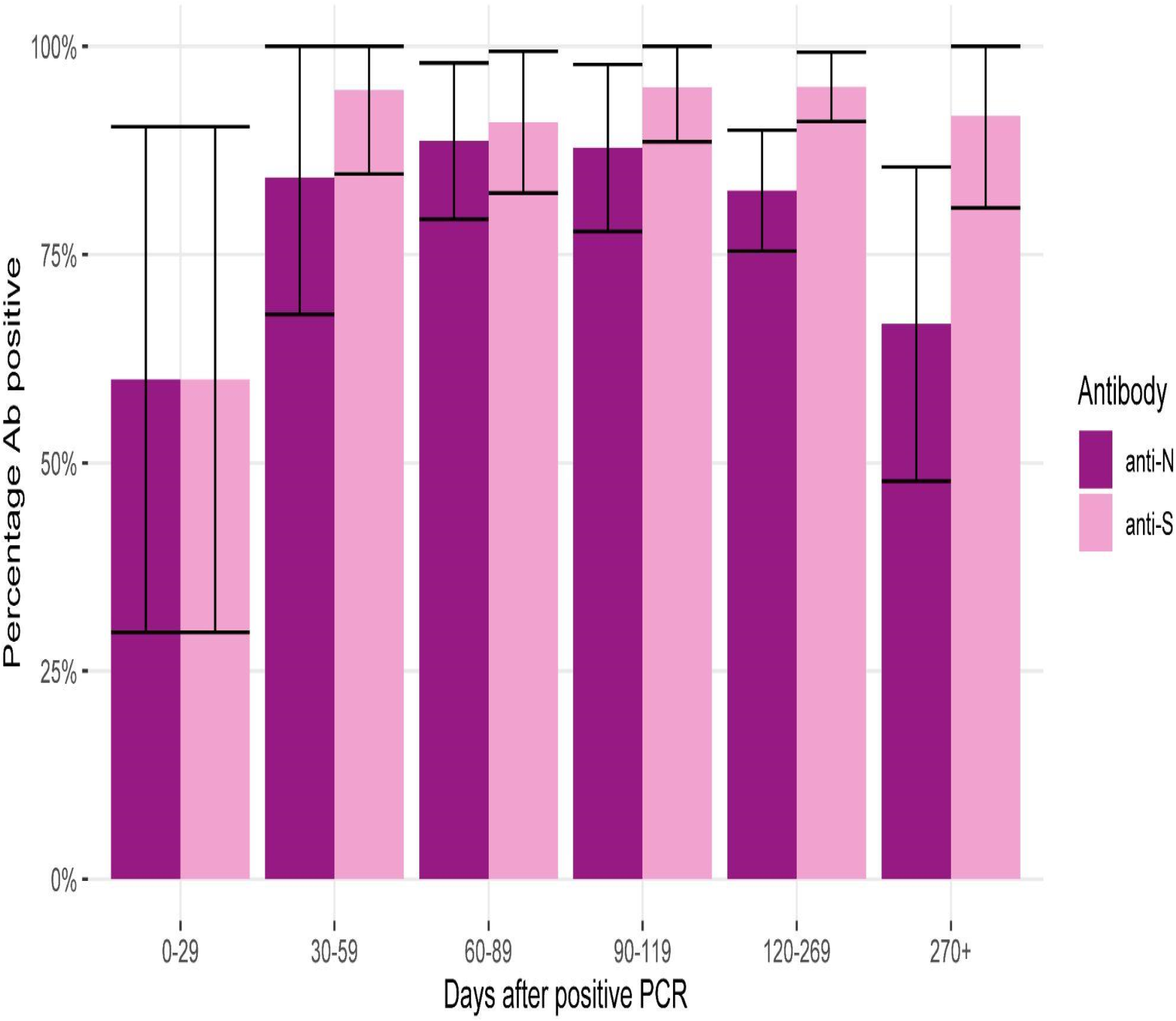
Percentage antibody positivity anti-N and anti-S among pre-vaccinated samples or samples taken from unvaccinated individuals, stratified by days post virological confirmed infection.

**Figure 5.**
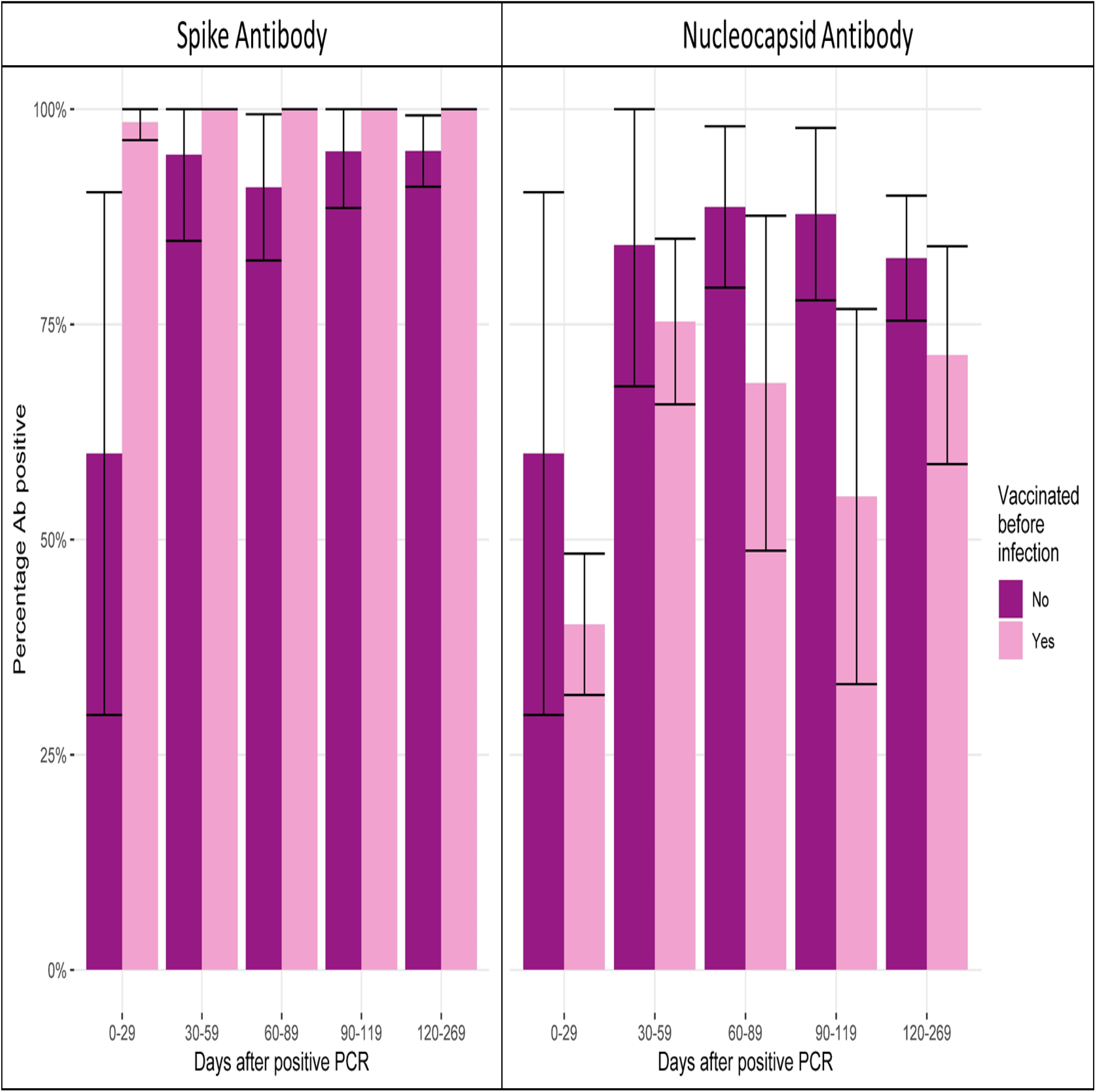
Proportion of antibody positivity for anti-S and anti-N by days post virological confirmed infection, stratified by vaccination prior to infection.

Anti-N positivity was generally higher for those with no prior vaccination (N = 244), compared to those with vaccination prior to infection (N = 305) with the most notable difference on days 90-119; 87.8% (95% CI 78-98) vs 55% (95% CI 33-77) p = 0.011. Anti-N titres from participants with no prior vaccination had higher median levels than in those with prior vaccination (39.6 COI vs 6.53 COI, p < 0.0001). The proportion of anti-S positivity was generally higher for samples taken from people with prior vaccination (N = 301) compared to those without prior vaccination (N = 244), with the most notable difference on days 0-29; 60% (95% CI 30-90) vs 98.5% (95% CI (96-100), p < 0.0001. The proportion of anti-S positivity was higher in those with vaccination only (N = 46,647), in comparison to infection only (N = 244), from 0 - 269 days. There were no results for 270+ days post vaccination and therefore were not included in this section of the analysis. The median age in the vaccination only group was higher than the infection only group (65 vs 43 years old, p < 0.0001).

## Discussion

We present the findings of a large community cohort study demonstrating anti-S and anti-N trends in participants with antibody results from 1 to 540 days since PCR confirmed infection. Our study found that approximately 4 out of 5 individuals were seropositive for anti-N at any point between 0 to 269 days after a PCR confirmed SARS-CoV-2 infection. Logistic regression models, both univariable and multivariable, only showed higher odds of a positive ant-N result for 35-49 year olds, compared to 18-34 year olds. The peak proportion in positivity for anti-N and median antibody level was 120-269 days and 90-119, respectively, after infection. Proportion of seropositivity and median antibody levels were higher among men than women from 120 days onwards, which was a similar pattern observed in ≥50 year olds compared to 18-49 year olds. Samples of those who were vaccinated prior to PCR confirmed infection, had a lower median anti-N and higher anti-S 30-269 days post infection. The proportion of anti-S positive samples were higher for vaccination only, compared to infection only, over time.

Previous studies have found that anti-N positivity after PCR confirmed infection ranges from 84.7% at 28 days to 68.2% at 293 days, with proportions being highest among those with severe symptom profiles. ^17,18^ Antibody positivity may be influenced by the viral load during infection, with higher viral loads causing higher levels of antigen exposure and more severe symptoms. ^19^ As our data is from a community cohort study it can be assumed that the symptom profiles may be less severe among our participants in comparison to hospital based longitudinal studies, and therefore leading to lower antibody levels.

Increasing age and sex in some studies have been associated with higher anti-S and anti-N IgG responses. ^18,20^ The only factor independently associated with seropositivity in the logistic regression model was age, with 35-49 year olds more likely to be seropositive than older groups, when compared to 18-34 year olds. Yet when assessing anti-N response over time, there was a difference between age categories 18-49 and 50+ on days 120-269 and 270+, with the former having an earlier peak and earlier antibody waning. Older people have a higher frequency of comorbidities, putting them at higher risk of severe disease, which may explain longer duration of anti-N positivity. ^21^ This being said, there was not a higher or lower odds of being seropositive if individuals have medical conditions associated with higher risk of COVID19 related mortality or taking immunosuppressant therapy. This is likely due to a small number of participants in these groups in our analyses. Although limited to 269 days post infection because of disparity in length of follow up between individuals, time interval between PCR and anti-N results was not accounted for in our models. Seropositivity has been shown to be affected by days since PCR, which may also explain why some differences were not found in our regression model. ^18^

The proportion of seropositive anti-N samples was above 80% from 30 days onwards, but only 42.6% of samples were positive between days 0-29. Although this is lower than other studies, where seroconversion rates have been 84.7% at 10-28 days respectively, these studies followed individuals who had been hospitalised due to COVID19. ^17^ When hospitalised individuals were stratified by symptom severity, the cumulative percentage of anti-N positive asymptomatic individuals was 60% at 22-28 days. ^17^ Other studies in hospitalised individuals have also demonstrated symptom severity is associated with an earlier peak in anti-N titres, as well as determining longer duration of anti-N positivity. ^22,23^ Difference in anti-N over time was seen when the data was disaggregated by sex, with earlier peak and waning in females. This may also be related to risk of disease severity, with a meta-analysis showing men are 2.41 times more at risk of developing severe disease in comparison to women. ^24^ Higher ACE2 expression, decreased B cell and NK cell-specific transcripts in men is suggested to be the cause of higher viral loads and therefore more severe symptoms. ^25,26^ Although we have not used symptom severity in our analysis, as Virus Watch is a community cohort study, it can be assumed that in comparison the symptom profiles are likely to be milder on average. This would explain a lower percentage of anti-N positive samples at 0-29 days, but provides a more accurate representation of anti-N longevity in the community setting.

After excluding post-vaccination samples, there was evidence of anti-N waning in both proportion of detectable anti-N in samples and median anti-N titres, which was not the case for anti-S. Earlier anti-N waning has also been observed in other studies, which monitored IgG. ^18,20^ A higher proportion of non-vaccinated samples were anti-N positive compared to samples from participants who had vaccination prior to infection. This trend was also reflected in median anti-N and ant-S titres. This finding is important when considering the use of anti-N as an alternative to anti-S for seroprevalence studies in highly vaccinated populations. The anti-S response seen between vaccinated and non-vaccinated is in keeping with the immunological mechanism of the vaccines, which induces an anti-S specific response. The proportion of anti-S positive results in the vaccination only cohort was higher at days 120-269 than anti-S response in the infection only group, indicating a more sustained anti-S response from vaccination. Although anti-S levels may differ between these groups, this is not necessarily an indication of risk of future infection. Comparison of infection risk between these two groups has provided conflicting information, with pooled results of randomised control trials showing no difference and observational studies favouring natural immunity. ^20^ Our results may be the outcome of confounding however, as the median age of the vaccine only group was higher than infection only.

The strengths of this study include a large sample size that spans various age groups and captures multiple underlying health conditions. We present serial antibody measurements using a highly sensitive, widely used validated commercial assay that provides quantitative readings. ^15^ There are limitations however, as a large proportion of the data is self-reported and therefore susceptible to reporting bias, as participants may only volunteer information they feel is relevant or necessary. Self-reporting is also susceptible to data entry errors. This led to some samples being excluded from any analyses based on erroneous reporting of vaccination status. Furthermore, a large proportion of the data in this analysis (e.g. medical conditions, medications) is only collected at registration, so did not account for changes in participant’s health or medications. The dates of PCR confirmed infection however, were SGSS data, allowing accurate calculation of time between infection and blood test. Those who did not have a positive PCR confirmed infection according to the linkage data were excluded from the analysis, therefore further reducing the sample size. The earliest positive PCR result was used in the analysis and subsequent PCR results were excluded. Therefore we are unable to report on subsequent asymptomatic reinfections/re-exposure which may boost antibody levels. ^19,27^

## Conclusion

As only 4 in 5 participants with prior PCR confirmed infection were anti-N positive at any time point up to 269 days after infection, seroprevalence studies on anti-N alone may underestimate the true cumulative incidence of infection. We have demonstrated a decline in anti-N levels from 120 days onwards, providing a better understanding of the limitations of seroprevalence studies. Duration of anti-N positivity is affected by age and sex; therefore serosurveillance may require shorter time window of testing post-infection.

## Data Availability

All data produced in the present study are available upon reasonable request to the authors

## Ethics

This study has been approved by the Hampstead NHS Health Research Authority Ethics Committee. Ethics approval number - 20/HRA/2320.

## Data availability

We aim to share aggregate data from this project on our website and via a “Findings so far” section on our website - https://ucl-virus-watch.net/. We will also be sharing individual record level data on a research data sharing service such as the Office of National Statistics Secure Research Service. In sharing the data we will work within the principles set out in the UKRI Guidance on best practice in the management of research data. Access to use of the data whilst research is being conducted will be managed by the Chief Investigators (ACH and RWA) in accordance with the principles set out in the UKRI guidance on best practice in the management of research data. We will put analysis code on publicly available repositories to enable their reuse.

