## Supplemental Tables and Figures for "Nucleocapsid and spike antibody responses post virologically confirmed SARS-CoV-2 infection: An observational analysis in the Virus Watch community cohort"

### Appendix: Summary tables

Table 1: Summary of medical conditions and medication variables from the baseline survey and which categories they were collapsed into for data analysis.

| Data collection variables | Data analysis variables |
|---|---|
| - Asthma<br>- Emphysema<br>- Chronic bronchitis<br>- Chronic Obstructive Pulmonary Disease<br>- Cystic fibrosis | Respiratory conditions |
| - Coronary heart disease<br>- Angina<br>- Heart attack or myocardial infarction | Ischaemic Heart Disease |
| - High blood pressure/hypertension | Hypertension |
| - Chronic kidney disease | Chronic Kidney Disease |
| - Insulin treated diabetes<br>- Other diabetes | Diabetes |
| - Coronary heart disease<br>- Angina<br>- Heart attack or myocardial infarction<br>- High blood pressure/hypertension<br>- Chronic kidney disease<br>- Insulin treated diabetes<br>- Other diabetes | Conditions associated with high mortality or intensive care admission (“COVID risk”) |
| - HIV | HIV |
| - Cancer or malignancy | Cancer (subdivided into haematological malignancy and non-haematological malignancy) |

| | |
|---|---|
| - Medication following an organ transplant<br>- Medicines such as steroid tablets that weaken the immune system<br>- Targeted therapy or chemotherapy for cancer treatment<br>- Radiotherapy for cancer treatment<br>- Other treatment or medication that may affect immune system | Immunosuppressant medications |

Table 2: Summary table of medical conditions for anti-N positive by all individuals with a PCR result and all individuals with a positive PCR result

| | **anti-N Positive (n / N)** | **Percentage Positive (%)** | **95% CI** |
|---|---|---|---|
| **Any Condition** | | | |
| No | 241 / 290 | 83.1% | (78.79, 87.42) |
| Yes | 150 / 193 | 77.72% | (71.85, 83.59) |
| Missing data | 41 / 54 | 75.93% | (64.52, 87.33) |
| **COVID Risk** | | | |
| No | 296 / 366 | 80.87% | (76.84, 84.9) |
| Yes | 91 / 113 | 80.53% | (73.23, 87.83 |
| Missing data | 45 / 58 | 77.59% | (66.85, 88.32) |

| | | | |
|---|---|---|---|
| **BMI** | | | |
| Normal BMI | 123 / 162 | 75.93% | (69.34, 82.51) |
| Obesity class 1 | 65 / 74 | 87.84% | (80.39, 95.29) |
| Obesity class 2 | 23 / 27 | 85.19% | (71.78, 98.59) |
| Obesity class 3 | 6 / 9 | 66.67% | (35.87, 97.47) |
| Pre-obese | 128 / 156 | 82.05% | (76.03, 88.07) |
| Missing data | 86 / 108 | 79.63% | (72.03, 87.23) |
| **Chronic Kidney Disease** | | | |
| No | 389 / 480 | 81.04% | (77.54, 84.55) |
| Yes | 2 / 3 | 66.67% | (13.32, 120.01) |
| Missing data | 41 / 54 | 75.93% | (64.52, 87.33) |
| **Diabetes** | | | |
| No | 365 / 455 | 80.22% | (76.56, 83.88) |

| | | | |
|---|---|---|---|
| Yes | 26 / 28 | 92.86% | (83.32, 102.4) |
| Missing data | 41 / 54 | 75.93% | (64.52, 87.33) |
| **Haematological Malignancy** | | | |
| No | 16 / 21 | 76.19% | (57.97, 94.41) |
| Missing data | 416 / 516 | 80.62% | (77.21, 84.03) |
| Missing data | 41 / 54 | 75.93% | (64.52, 87.33) |
| **Hypertension** | | | |
| No | 321 / 393 | 81.68% | (77.85, 85.5) |
| Yes | 70 / 90 | 77.78% | (69.19, 86.37) |
| Missing data | 41 / 54 | 75.93% | (64.52, 87.33) |
| **Ischaemic Heart Disease** | | | |
| No | 379 / 467 | 81.16% | (77.61, 84.7) |
| Yes | 12 / 16 | 75% | (53.78, 96.22) |

| | | | |
|---|---|---|---|
| Missing data | 41 / 54 | 75.93% | (64.52, 87.33) |
| **Malignancy** | | | |
| No | 375 / 462 | 81.17% | (77.6, 84.73) |
| Yes | 16 / 21 | 76.19% | (57.97, 94.41) |
| Missing data | 41 / 54 | 75.93% | (64.52, 87.33) |
| **Non-Haematological Malignancy** | | | |
| No | 16 / 21 | 76.19% | (57.97, 94.41) |
| Missing data | 416 / 516 | 80.62% | (77.21, 84.03) |
| **Respiratory Conditions** | | | |
| No | 322 / 393 | 81.93% | (78.13, 85.74) |
| Yes | 69 / 90 | 76.67% | (67.93, 85.41) |
| Missing data | 41 / 54 | 75.93% | (64.52, 87.33) |

Table 3: Summary table of immunotherapy for anti-N positive by all individuals with a PCR result and all individuals with a positive PCR result

| | anti-N Positive (n / N) | Percentage Positive (%) | 95% CI |
|---|---|---|---|
| **Any immunosuppressant** | | | |
| No | 223 / 276 | 80.8% | (76.15, 85.44) |
| Yes | 13 / 19 | 68.42% | (47.52, 89.32) |
| Missing data | 196 / 242 | 80.99% | (76.05, 85.94) |
| **Chemotherapy/Cancer treatment** | | | |
| No | 234 / 293 | 79.86% | (75.27, 84.45) |
| Yes | 2 / 2 | 100% | (100, 100) |
| Missing data | 196 / 242 | 80.99% | (76.05, 85.94) |
| **Steroids** | | | |
| No | 232 / 288 | 80.56% | (75.99, 85.13) |

| | | | |
|---|---|---|---|
| Yes | 4 / 7 | 57.14% | (20.48, 93.8) |
| Missing data | 196 / 242 | 80.99% | (76.05, 85.94) |
| **Transplant medication** | | | |
| No | 236 / 294 | 80.27% | (75.72, 84.82) |
| Yes | 0 / 1 | 0% | (0, 0) |
| Missing data | 196 / 242 | 80.99% | (76.05, 85.94) |

Table 4: Logistic Regression model for odds ratio of being anti-N positive at any time point after PCR confirmed infection

| | **Univariable Logistic Regression** | | | **Multivariable Logistic Regression** | | | **Median Days Post PCR** | |
|---|---|---|---|---|---|---|---|---|
| | **OR** | **95 CI** | **p-value** | **OR** | **95 CI** | **p-value** | **Negative anti-N** | **Positive anti-N** |
| **Age category** | | | | | | | | |
| 18-34 | 1 | 1 | 1 | 1 | 1 | 1 | 53 | 73 |
| 35-49 | 1.99 | (1.01-3.92) | 0.05 | 2.02 | (1.02-3.98) | 0.04 | 43 | 86 |
| 50-64 | 1.3 | (0.7-2.37) | 0.39 | 1.31 | (0.71-2.39) | 0.38 | 29.5 | 82 |

| | | | | | | | | |
|---|---|---|---|---|---|---|---|---|
| 65-79 | 1.81 | (0.9-3.64) | 0.09 | 1.78 | (0.89-3.58) | 0.1 | 42.5 | 90 |
| **Sex** | | | | | | | | |
| Female | 1 | 1 | 1 | 1 | 1 | 1 | 46 | 80 |
| Male | 1.46 | (0.94-2.32) | 0.1 | 1.45 | (0.93-2.3) | 0.11 | 20 | 94 |
| **Obesity** | | | | | | | | |
| No | 1 | 1 | 1 | 1 | 1 | 1 | 40 | 78 |
| Yes | 1.57 | (0.88-2.93) | 0.14 | 1.68 | (0.94-3.16) | 0.09 | 35 | 91 |
| **COVID risk** | | | | | | | | |
| No | 1 | 1 | 1 | 1 | 1 | 1 | 42 | 80 |

| | | | | | | | | |
|---|---|---|---|---|---|---|---|---|
| Yes | 0.98 | (0.58-1.7) | 0.94 | 0.88 | (0.5-1.58) | 0.66 | 28.5 | 86 |
| **Any immunosuppressant** | | | | | | | | |
| No | 1 | 1 | 1 | 1 | 1 | 1 | 42 | 81 |
| Yes | 0.51 | (0.19-1.52) | 0.2 | 0.53 | (0.2-1.56) | 0.22 | 9.5 | 50 |

Table 5: Summary of the monthly antibody testing cohort, with valid samples, and the entire Virus Watch Cohort

| | **Virus Watch Cohort (N = 50773)** | **Antibody Testing Cohort (N = 13802)** | **Individual Level Analysis (N = 537)** |
|---|---|---|---|
| **Age category** | | | |
| 18-34 | 7301 (14.38) | 973 (7.05) | 79 (14.71) |
| 35-49 | 9148 (18.02) | 1983 (14.37) | 143 (26.63) |
| 50-64 | 13496 (26.58) | 4819 (34.92) | 193 (35.94) |
| 65-79 | 12853 (25.31) | 5717 (41.42) | 120 (22.35) |
| 80+ | 960 (1.89) | 288 (2.09) | 2 (0.37) |
| Missing data | 7015 (13.82) | 22 (0.16) | 0 (0) |
| **Sex** | | | |
| Female | 23431 (46.15) | 7853 (56.9) | 324 (60.34) |
| Male | 18885 (37.19) | 5906 (42.79) | 211 (39.29) |
| Missing data | 8457 (16.66) | 43 (0.31) | 2 (0.37) |

| | | | |
|---|---|---|---|
| **Ethnicity** | | | |
| Black | 469 (0.92) | 57 (0.41) | 5 (0.93) |
| Mixed Categories | 624 (1.23) | 77 (0.56) | 6 (1.12) |
| Other Asian | 397 (0.78) | 81 (0.59) | 2 (0.37) |
| Other/missing | 8992 (17.71) | 79 (0.57) | 8 (1.49) |
| South Asian | 2687 (5.29) | 219 (1.59) | 16 (2.98) |
| White | 37337 (73.54) | 13242 (95.94) | 500 (93.11) |
| Missing data | 267 (0.53) | 47 (0.34) | |
| **Any Condition** | | | |
| No | 21697 (42.73) | 6582 (47.69) | 290 (54) |
| Yes | 14596 (28.75) | 5978 (43.31) | 193 (35.94) |
| Missing data | 14480 (28.52) | 1242 (9) | 54 (10.06) |
| **BMI** | | | |

| | | | |
|---|---|---|---|
| Normal | 11581 (22.81) | 4802 (34.79) | 162 (30.17) |
| Obesity class 1 | 4214 (8.3) | 1688 (12.23) | 74 (13.78) |
| Obesity class 2 | 1428 (2.81) | 546 (3.96) | 27 (5.03) |
| Obesity class 3 | 721 (1.42) | 268 (1.94) | 9 (1.68) |
| Pre-obese | 9897 (19.49) | 4217 (30.55) | 156 (29.05) |
| Underweight | 425 (0.84) | 132 (0.96) | 1 (0.19) |
| Missing data | 22507 (44.33) | 2149 (15.57) | 108 (20.11) |
| **Chronic Kidney Disease** | | | |
| No | 35922 (70.75) | 12392 (89.78) | 480 (89.39) |
| Yes | 371 (0.73) | 168 (1.22) | 3 (0.56) |
| Missing data | 14480 (28.52) | 1242 (9) | 54 (10.06) |
| **COVID Risk** | | | |
| No | 27242 (53.65) | 8543 (61.9) | 366 (68.16) |

| | | | |
|---|---|---|---|
| Yes | 9051 (17.83) | 4017 (29.1) | 113 (21.04) |
| Missing data | 14480 (28.52) | 1242 (9) | 58 (10.8) |
| **Diabetes** | | | |
| No | 33969 (66.9) | 11666 (84.52) | 455 (84.73) |
| Yes | 2324 (4.58) | 894 (6.48) | 28 (5.21) |
| Missing data | 14480 (28.52) | 1242 (9) | 54 (10.06) |
| **Haematological Malignancy** | | | |
| No | 2073 (4.08) | 980 (7.1) | 21 (3.91) |
| Yes | 217 (0.43) | 96 (0.7) | |
| Missing data | 48483 (95.49) | 12726 (92.2) | 516 (96.09) |
| **HIV** | | | |
| No | 36200 (71.3) | 12538 (90.84) | 481 (89.57) |
| Yes | 93 (0.18) | 22 (0.16) | 2 (0.37) |

| | | | |
|---|---|---|---|
| Missing data | 14480 (28.52) | 1242 (9) | 54 (10.06) |
| **Hypertension** | | | |
| No | 29063 (57.24) | 9252 (67.03) | 393 (73.18) |
| Yes | 7230 (14.24) | 3308 (23.97) | 90 (16.76) |
| Missing data | 14480 (28.52) | 1242 (9) | 54 (10.06) |
| **Ischaemic Heart Disease** | | | |
| No | 34927 (68.51) | 11968 (86.71) | 467 (86.96) |
| Yes | 1366 (2.68) | 592 (4.29) | 16 (2.98) |
| Missing data | 14480 (28.52) | 1242 (9) | 54 (10.06) |
| **Malignancy** | | | |
| No | 33996 (66.69) | 11484 (83.21) | 462 (86.03) |
| Yes | 2297 (4.51) | 1076 (7.8) | 21 (3.91) |
| Missing data | 14480 (28.52) | 1242 (9) | 54 (10.06) |

| | | | |
|---|---|---|---|
| **Non-Haematological Malignancy** | | | |
| No | 184 (0.36) | 86 (0.62) | 0 (0) |
| Yes | 2106 (4.15) | 990 (7.17) | 21 (3.91) |
| Missing data | 48483 (95.49) | 12726 (92.2) | 516 (96.09) |
| **Respiratory Conditions** | | | |
| No | 30161 (59.16) | 10389 (75.27) | 393 (73.18) |
| Yes | 6132 (12.03) | 2171 (15.73) | 90 (16.76) |
| Missing data | 14480 (28.52) | 1242 (9) | 54 (10.06) |
| **Any immunosuppressant** | | | |
| No | 18367 (36.17) | 7649 (55.42) | 276 (51.4) |
| Yes | 1861 (3.67) | 736 (5.33) | 19 (3.54) |
| Missing data | 30545 (60.16) | 5417 (39.25) | 242 (45.07) |
| **Steroids** | | | |

| | | | |
|---|---|---|---|
| No | 19599 (38.6) | 8119 (58.82) | 288 (53.63) |
| Yes | 629 (1.24) | 266 (1.93) | 7 (1.3) |
| Missing data | 30545 (60.16) | 5417 (39.25) | 242 (45.07) |
| **Chemotherapy/Cancer treatment** | | | |
| No | 20019 (39.43) | 8295 (60.1) | 293 (54.56) |
| Yes | 209 (0.41) | 90 (0.65) | 2 (0.37) |
| Missing data | 30545 (60.16) | 5417 (39.25) | 242 (45.07) |
| **Transplant medication** | | | |
| No | 20164 (39.71) | 8356 (60.54) | 294 (54.75) |
| Yes | 64 (0.13) | 29 (0.21) | 1 (0.19) |
| Missing data | 30545 (60.16) | 5417 (39.25) | 242 (45.07) |

Table 6: Power calculations for univariable logistic regression model for binary exposure variables

| Exposure | Significance level | Power | Sample size | Pr(x=1) | Pr(y = 1 \| x=0) | Detectable/alternative OR |
|---|---|---|---|---|---|---|
| Male sex | 0.05 | 0.8 | 535 | 0.39 | 0.47 | 1.653 |
| Obesity | 0.05 | 0.8 | 272 | 0.40 | 0.45 | 2.031 |
| COVID risk | 0.05 | 0.8 | 479 | 0.24 | 0.62 | 1.985 |
| Immunosuppressant | 0.05 | 0.8 | 295 | 0.06 | 0.76 | NaN* |

*Need a sample size of 900 for in order to calculate a detectable OR. Sample size of 900 produces a detectable OR of 5.35

Calculations using algorithms described in Demidenko E. (2007). "Sample size determination for logistic regression revisited." Statistics in Medicine 26:3385-3397 and Demidenko E. (2008) "Sample size and optimal design for logistic regression with binary interaction." Statistics in Medicine, 27:36-46. Link to online calculator can be found here: https://www.dartmouth.edu/~eugened/power-samplesize.php

### Appendix: Figures

Figure 1: Proportion of samples positive for anti-S separated into infection only and vaccination only groups, over time since infection/first vaccination.

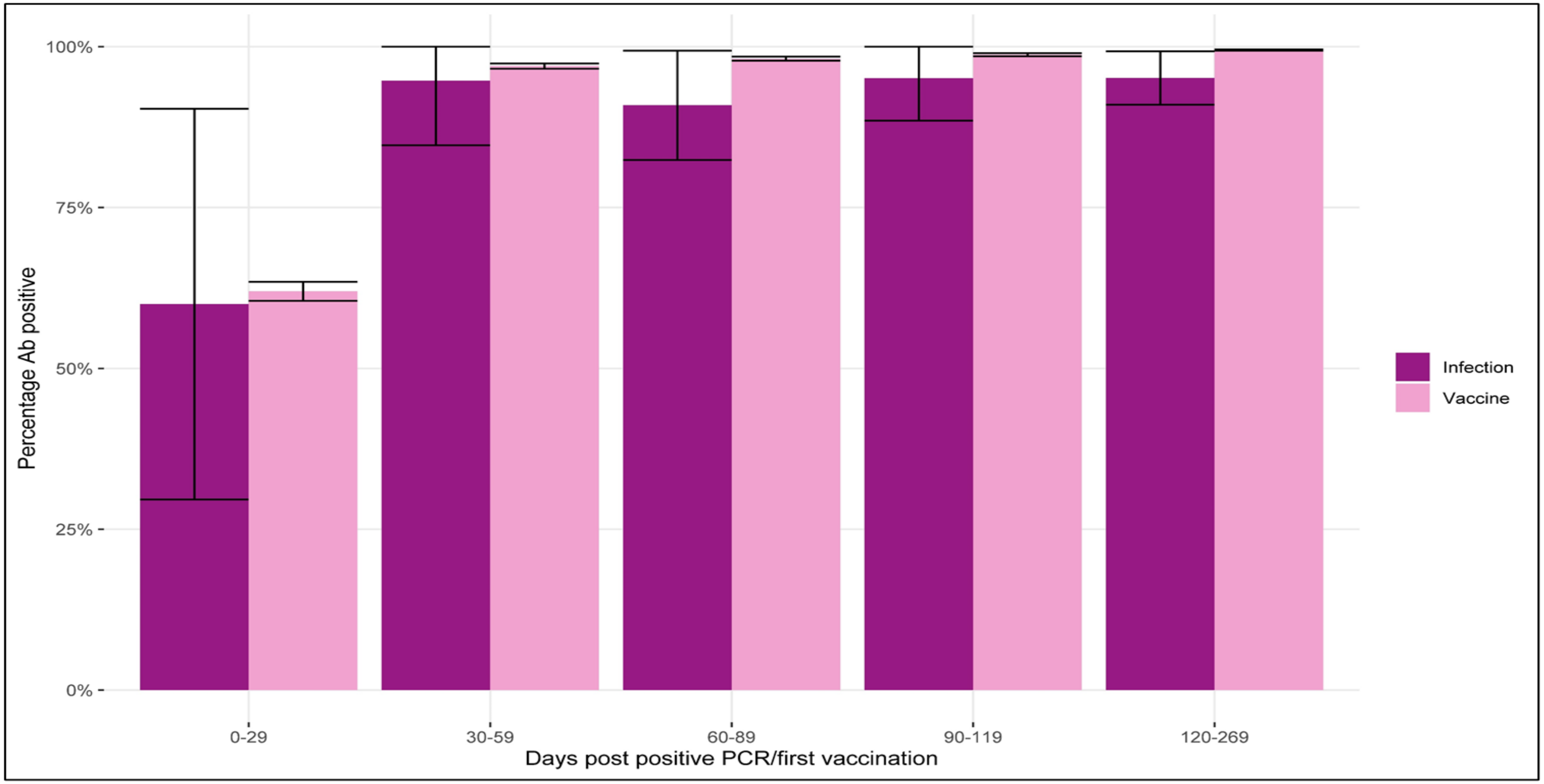

100%
75%
50%
25%
0%
Percentage Ab positive
0-29
30-59
60-89
90-119
120-269
Days post positive PCR/first vaccination
Infection
Vaccine
